## Supplementary Material for "Quantitative Multi-pathway Assessment of Exposure to Fecal Contamination for Infants in Rural Ethiopia"

### Supplementary Materials

#### A Supplementary Tables

Supplementary Table 1. Lab methods for different sample types.

<table>
<tr><th>Sample Type</th><th>Collected Volume</th><th>Detection Method</th><th>Detection Volume</th></tr>
<tr><td>Areola Swab</td><td>4 mL</td><td rowspan="2">EC MUG</td><td rowspan="2">5 wells of 180 uL</td></tr>
<tr><td>Fomites</td><td>Sponge (5 mL)</td></tr>
<tr><td>Handrinse</td><td>200 mL</td><td rowspan="3">EC MUG</td><td rowspan="3">88 wells of 180 uL</td></tr>
<tr><td>Bathing Water</td><td>1 L</td></tr>
<tr><td>Drinking Water</td><td>1 L</td></tr>
<tr><td>Food</td><td>1–10 g</td><td>EC MUG/Chromocult</td><td>5 wells of 180 uL</td></tr>
<tr><td>Breast Milk</td><td>0.5–5 mL</td><td rowspan="2">Chromocult</td><td rowspan="2">200 uL with serial dilutions</td></tr>
<tr><td>Soil</td><td>Boot socks (15 mL)</td></tr>
</table>

Supplementary Table 2. Estimated parameters of normal distributions for log 10 scale *E. coli* concentrations at two timepoints.

| | Timepoint 1 | | Timepoint 2 | | Total | | |
|---|---|---|---|---|---|---|---|
| Sample Type | Mean | SD | Mean | SD | Mean | SD | Unit |
| Areola Swab | -0.043 | 0.674 | -0.434 | 1.141 | -0.287 | 0.945 | per swab |
| Breast Milk | -0.203 | 1.103 | -1.573 | 1.197 | -0.822 | 1.218 | per mL |
| Mother Handrinse | 2.051 | 1.070 | 1.947 | 0.981 | 2.000 | 1.029 | per pair of hands |
| Sibling Handrinse | 2.029 | 0.926 | 2.001 | 0.924 | 2.020 | 0.925 | per pair of hands |
| Infant Handrinse | 1.145 | 0.941 | 1.885 | 1.074 | 1.490 | 1.101 | per pair of hands |
| Bathing Water | -0.324 | 1.093 | -0.420 | 0.975 | -0.371 | 1.045 | per mL |
| Drinking Water | -0.709 | 0.727 | -1.176 | 0.714 | -0.944 | 0.758 | per mL |
| Fomite | 0.765 | 0.864 | 0.680 | 1.483 | 0.725 | 1.210 | per sponge |
| Food | -0.012 | 1.570 | -0.658 | 1.681 | -0.436 | 1.663 | per gram |
| Soil | | | 5.188 | 0.866 | 5.188 | 0.866 | per boot sock |

### B Supplementary Figure Captions

Supplementary Figure 1. Proportion of time spent for duration-based behaviors by Timepoint. Boxplots show distributions of the proportions of time spent on specific activities, compartments, or locations among infants. The central line in the box indicates the median value, while the box limits indicate the first and third quartile. The points outside of the box indicate outliers.

Supplementary Figure 2. Rate (times per hour) of frequency-based behaviors by Timepoint. Boxplots show distributions of the rates of frequency-based behaviors among infants. The central line in the box indicates the median value, while the box limits indicate the first and third quartile. The points outside of the box indicate outliers.

Supplementary Figure 3. Observed activity sequences at Timepoint 1. The morning and afternoon sessions were combined for each infant. The x-axis shows the time and the y-axis shows the age of infants, which is sorted in ascending order (from bottom to top). “NA” represents the time period not observed.

Supplementary Figure 4. Observed activity sequences at Timepoint 2. The morning and afternoon sessions were combined for each infant. The x-axis shows the time and the y-axis shows the age of infants, which is sorted in ascending order (from bottom to top). “NA” represents the time period not observed.

Supplementary Figure 5. Observed compartment sequences at Timepoint 1. The morning and afternoon sessions were combined for each infant. The x-axis shows the time and the y-axis shows the age of infants, which is sorted in ascending order (from bottom to top). “NA” represents the time period not observed.

Supplementary Figure 6. Observed compartment sequences at Timepoint 2. The morning and afternoon sessions were combined for each infant. The x-axis shows the time and the y-axis shows the age of infants, which is sorted in ascending order (from bottom to top). “NA” represents the time period not observed.

Supplementary Figure 7. (a) Observed behavior transition network at Timepoint 1 (6050 transitions); (b) observed behavior transition network at Timepoint 2 (6568 transitions). The text inside the node shows the activity. The color of the node represents the compartment: light gray is Carried by Mother; dark gray is Carried by Others; pink is Down on a Surface with Barriers; red is Down on the Bare Ground. For the location, the nodes above the horizontal line are within homestead, while those below the horizontal line are out of homestead. Arrows indicate transitions between states (i.e., combinations of activity, compartment, and location), and strengths (numbers of times the transition was observed) indicated by arrow width and shade (darker arrows indicate higher frequency).

Supplementary Figure 8. Estimated rate (times per hour) of frequency-based behaviors conditional on the state at Timepoint 1. The color represents the rate: darker colors show higher rates.

Supplementary Figure 9. Estimated rate (times per hour) of frequency-based behaviors conditional on the state at Timepoint 2. The color represents the rate: darker colors show higher rates.

Supplementary Figure 10. Correlation matrix for log10 scale *E. coli* concentration levels between different sample types. In the correlation matrix, plots in the bottom left half show the scatterplots of log10 scale *E. coli* concentration levels for pairs of sample types. Plots on the diagonal line show the histogram of log10 scale *E. coli* concentration levels by sample type. Plots on the top right half show the Spearman correlation coefficients of log10 scale *E. coli* concentration levels between different sample types. The stars in the figure define the level of significance. * = 0.05, ** = 0.01, *** = 0.001.

Supplementary Figure 11. Exposure to *E. coli* by source and timepoint. The bar charts show the fraction of simulated days that children are exposed, and boxplots show the estimated daily dose of *E. coli* ($log_{10}$ CFU/day).
